# Is Vitamin D Deficiency Linked to Depression? Insights from a Nationally Representative US Sample

**DOI:** 10.1101/2025.05.12.25327452

**Authors:** Sonia Mehta, Nikhila Chelikam, Shreya Shamhavi, Jamarc Simon, Pramil Cheriyath, Vinod Nookala

## Abstract

**Background:** Vitamin D deficiency (VDD) has been known to be involved in neuropsychiatric conditions, including depression. However, the association between VDD and specific depressive symptoms remains unclear. This study aims to investigate the relationship between VDD and individual depressive symptoms using data from a large, nationally representative U.S. population.

**Methods:** We conducted a retrospective cross-sectional analysis of 10,914 adults from the 2007 to 2018 NHANES cycles with available serum 25(OH)D levels and complete PHQ-9 derived Depression Screener (DPQ) data. VDD was defined as <30 ng/mL. Specific depressive symptoms like little interest, feeling depressed, low energy, negative self-image, and suicidal ideation were assessed. Multivariable logistic regression was performed after adjusting for demographics and comorbidities.

**Results:** VDD was present in 8% of participants and was more prevalent among younger adults, females, and individuals with obesity. Key depressive symptoms such as low interest (29.9% vs. 24.8%), feeling down (30.4% vs. 24.5%), and fatigue (55.0% vs. 48.9%) were significantly more common in the VDD group (p < 0.01 for all). VDD was independently associated with increased odds of any depressive symptoms (adjusted odds ratio [aOR] 1.18; 95% CI: 1.02-1.36; p = 0.028). Female gender (aOR 1.78; 95% CI: 1.65-1.92; p <0.001), age <60 (aOR 1.49; 95% CI: 1.36-1.63; p <0.001), obesity (aOR 1.21; 95% CI: 1.11-1.32; p <0.001), diabetes (aOR 1.32; 95% CI: 1.16-1.49; p <0.001), hypertension (aOR 1.25; 95% CI: 1.14-1.37; p < 0.001), and high triglycerides (aOR 1.19; 95% CI: 1.08-1.30; p = 0.0003) were stronger predictors of depression.

**Conclusion:** VDD is modestly but significantly associated with depressive symptoms, particularly little interest and low energy. Although the effect size is smaller than that of metabolic and demographic factors, VDD may represent a modifiable target in depression prevention and management strategies.

## Introduction

Vitamin D plays a crucial role in calcium homeostasis, immune modulation, and neuroprotection. Beyond its well-known effects on bone health, emerging research has linked vitamin D deficiency (VDD) to neurological and psychiatric disorders, including depression (Holick, 2007; Eyles et al., 2005). Major depressive disorder is a significant global health burden and is projected by the World Health Organization (WHO) to become the leading cause of disease burden by 2030 (Malhi & Mann, 2018). Depression is nearly twice as prevalent in women, likely due to biological, hormonal, and psychosocial factors (Pedersen et al., 2014; Drevets et al., 2008).

Several observational studies and meta-analyses suggest a link between VDD and depressive symptoms, though causality remains uncertain due to confounding factors (Anglin et al., 2013; Mulugeta et al., 2020). For instance, a meta-analysis (Anglin et al.,2013) reported a significant association between low vitamin D levels and depression, while a recent study (dos Santos et al.,2024) from the Brazilian Longitudinal Study of Aging (ELSI-Brazil) found higher depressive symptoms among individuals with insufficient vitamin D levels, supporting a possible biological relationship. However, inconsistencies in findings across studies may stem from variations in study design, population characteristics, and definitions of VDD and depression (Kerr et al., 2015; Kim et al., 2020).

In this study, we investigate the association between VDD and specific depressive symptoms, including loss of interest, depressed mood, low energy, negative self-image, and suicidal ideation, using a large, nationally representative sample. We aim to clarify whether VDD contributes independently to depressive symptoms, with the hope that findings may inform public health strategies and clinical approaches, including targeted vitamin D optimization for at-risk populations.

## Materials and Methods

### Data Source

Data were obtained from the National Health and Nutrition Examination Survey (NHANES), a cross-sectional, nationally representative survey designed to assess the health and nutritional status of the U.S. population. NHANES is conducted by the Centers for Disease Control and Prevention (CDC) and employs a multistage probability sampling strategy to ensure representativeness. Every year, NHANES experts assess 5,000 individuals across the United States in mobile examination centers. The National Center for Health Statistics (NCHS) Research Ethics Review Board approves the study design and protocols annually. The NHANES dataset integrates demographic, socioeconomic, dietary, laboratory, and health-related data and a physical examination performed by trained medical professionals. All NHANES data are publicly available on the CDC website (https://www.n.cdc.gov/nchs/nhanes/Default.aspx).

### Study Population and Definitions

This study was a retrospective cross-sectional analysis of NHANES data collected between 2007 and 2018. Individual datasets from each NHANES cycle were downloaded and merged using Statistical Analysis System (SAS) Version 9.4.

### Inclusion and Exclusion Criteria

Participants with available serum 25-hydroxyvitamin D [25(OH)D] levels (LBXVIDMS) and complete DPQ responses were included. Participants were excluded if they had missing values for vitamin D levels and responses to DPQ questions.

#### Vitamin D Classification

Vitamin D levels were categorized as deficient (<30 ng/mL) and sufficient (≥30 ng/mL).

#### Depressive Symptoms Assessment

Depressive symptoms were assessed using the NHANES Depression Screener (DPQ), derived from the PHQ-9. The following DPQ questions were analyzed:

- Little Interest or Pleasure in Doing Things (DPQ010)
- Feeling Down, Depressed, or Hopeless (DPQ020)
- Feeling Tired or Having Little Energy (DPQ040)
- Feeling Bad About Oneself or a Failure (DPQ060)
- Thoughts of Being Better Off Dead or Self-Harm (DPQ090)

Participants were categorized into four age groups (RIDAGEYR): up to 19 years (<20 years), 20 – 39 years, 40–59 years, and 60+ years (≥60 years). Gender (RIAGENDR) was classified as Male or Female. Race/ethnicity (RIDRETH1) was grouped into Hispanic, non-Hispanic, and Other. Education level (DMDEDUC2) was categorized into Less than High School, High School, and More than High School.

Comorbidities were defined as follows: Diabetes was identified by a “Yes” response to DIQ010, and Hypertension was determined by a “Yes” response to BPQ020. Body Mass Index (BMI) was classified as Normal (<25 kg/m^2^), Overweight (25–29.9 kg/m^2^), and Obese (≥30 kg/m^2^). Alcohol use was categorized as Yes (ALQ101 = 1) or No (ALQ101 = 2).

Triglyceride (TGL) levels were classified as High (≥150 mg/dL) or Normal (<150 mg/dL). Total cholesterol levels were categorized as Desirable (<200 mg/dL), Borderline High (200–239 mg/dL), and High (≥240 mg/dL).

## Statistical Analysis

Descriptive statistics and chi-square tests were conducted to compare depressive symptom prevalence between vitamin D-deficient and sufficient individuals. Multivariable logistic regression models estimated odds ratios (ORs) and 95% confidence intervals (CIs), adjusting for demographics and comorbidities. Statistical significance was set at p<0.05. SAS Version 9.4 was used for all analyses.

## Results

### Demographic Characteristics of Participants by Vitamin D Status

Of the 10,914 respondents included in the analysis, 882 individuals (approximately 8%) were classified as vitamin D deficient (VDD). Vitamin D deficiency was more prevalent in younger adults, particularly those aged 20–39 years, who made up 43.2% of the VDD group compared to 31.9% in the vitamin D sufficient (VDS) group (p < 0.0001). In contrast, those over 60 years comprised only 23.4% of the VDD group, whereas they made up 34.8% of the VDS group. Gender distribution also differed significantly by vitamin D status. Females accounted for 55% of the VDD group compared to 50.4% of the VDS group (p = 0.0094). Although racial/ethnic distribution did not reach statistical significance (p = 0.1075), there was a trend toward fewer Hispanics in the VDD group (23.7% vs. 26.8%) and more non-Hispanic individuals (66.4% vs. 63.1%).

Educational attainment showed marked variation: 71% of VDD individuals had more than a high school education, compared to 75.3% in the VDS group, while the proportion with only a high school education was higher in the VDD group (20% vs. 14.2%; p < 0.0001). (Table 1).

**Table 1:** Demographic Characteristics.

|  | Vitamin D Deficient |  | Vitamin D Sufficient |  | *p_value* |
| --- | --- | --- | --- | --- | --- |
| Total number of respondents | 882 |  | 10032 |  |  |
| Age Category | | | | | $< 0.0001$ |
| 20-39 years | 381 | 43.20% | 3204 | 31.94% |  |
| 40-59 years | 295 | 33.45% | 3335 | 33.24% |  |
| > 60 years | 206 | 23.36% | 3493 | 34.82% |  |
| Gender |  |  |  |  | 0.0094 |
| Female | 485 | 54.99% | 5059 | 50.43% |  |
| Male | 397 | 45.01% | 4973 | 49.57% |  |
| Race/Ethnicity |  |  |  |  | 0.1075 |
| Hispanic | 209 | 23.70% | 2688 | 26.79% |  |
| Non-Hispanic | 586 | 66.44% | 6326 | 63.06% |  |
| Other | 87 | 9.86% | 1018 | 10.15% |  |
| Education Level |  |  |  |  | <0.0001 |
| <High School | 80 | 9.07% | 1048 | 10.45% |  |
| High School | 176 | 19.95% | 1426 | 14.21% |  |
| >High School | 626 | 70.98% | 7558 | 75.34% |  |

### Prevalence of Clinical Risk Factors in Vitamin D Deficient vs. Sufficient Individuals

Obesity was significantly more common among those with VDD: 51.3% had a BMI ≥30 kg/m^2^ compared to 35.5% of vitamin D sufficient individuals (p < 0.0001). Conversely, the proportion of participants with normal BMI was lower in the VDD group (23.8% vs. 30.4%). Interestingly, high total cholesterol was *less prevalent* in the VDD group (10.1% vs. 12.8%, p = 0.0222), and high triglycerides showed a non-significant trend (23.1% vs. 25.8%, p = 0.0862). There were no significant differences in the prevalence of diabetes (14.7% vs. 13.2%, p = 0.1847), hypertension (38.4% vs. 36.7%, p = 0.3064), or alcohol use (70.6% vs. 72.1%, p = 0.3666) between groups. (Table 2).

**Table 2:** Prevalence of Risk Factors.

|  | Vitamin D Deficient |  | Vitamin D Sufficient |  | *p_value* |
| --- | --- | --- | --- | --- | --- |
| Total number of respondents | 882 |  | 10032 |  |  |
| BMI |  |  |  |  | <0.0001 |
| Normal (<25 kg/m<sup>2</sup>) | 210 | 23.81% | 3051 | 30.41% |  |
| Overweight (25-29.9 kg/m<sup>2</sup>) | 220 | 24.94% | 3419 | 34.08% |  |
| Obese (>=30 kg/m<sup>2</sup>) | 452 | 51.25% | 3562 | 35.51% |  |
| Diabetes | 130 | 14.74% | 1320 | 13.16% | 0.1847 |
| Hypertension | 339 | 38.44% | 3682 | 36.70% | 0.3064 |
| High Triglycerides | 204 | 23.13% | 2584 | 25.76% | 0.0862 |
| High Total Cholesterol | 89 | 10.09% | 1279 | 12.75% | 0.0222 |
| Alcohol Use | 623 | 70.63% | 7229 | 72.06% | 0.3666 |

### Prevalence of Specific Depressive Symptoms by Vitamin D Status

Individuals with vitamin D deficiency reported a higher prevalence of key depressive symptoms, including little interest (29.9% vs. 24.8%, p = 0.0008), feeling down (30.4% vs. 24.5%, p = 0.0001), and low energy (55.0% vs. 48.9%, p = 0.0006), compared to those with sufficient vitamin D. However, differences in negative self-image (19.2% vs. 17.0%, p = 0.0972) and suicidal thoughts (4.0% vs. 3.6%, p = 0.5734) were not statistically significant. (Table 3)

**Table 3:** Association between Vitamin D Deficient and specific depressive symptoms.

|  | Vitamin D Deficient |  | Vitamin D Sufficient |  | *p_value* |
| --- | --- | --- | --- | --- | --- |
| Total number of respondents | 882 |  | 10032 |  |  |
| Little Interest | 264 | 29.93% | 2490 | 24.82% | 0.0008 |
| Feeling Depressed | 268 | 30.39% | 2458 | 24.50% | 0.0001 |
| Low Energy | 485 | 54.99% | 4910 | 48.94% | 0.0006 |
| Negative Self-Image | 169 | 19.16% | 1702 | 16.97% | 0.0972 |
| Suicidal Thoughts | 35 | 3.97% | 361 | 3.60% | 0.5734 |

### Multivariable Logistic Regression Predicting Depressive Symptoms

In the multivariable logistic regression analysis, vitamin D deficiency remained a significant predictor of depressive symptoms even after adjusting for age, gender, BMI, comorbidities, and other covariates. Individuals with VDD had an 18% higher likelihood of reporting depressive symptoms (aOR 1.18, 95% CI: 1.02–1.36, p = 0.0281).

Female gender showed the strongest association with depression, with women nearly twice as likely to report symptoms compared to men (aOR 1.78, 95% CI: 1.65–1.92, p < 0.0001). Age was also a significant factor; adults under 60 years had higher odds of depression compared to older adults (aOR 1.49, 95% CI: 1.36–1.63, p < 0.0001).

Obesity independently predicted depression, with an adjusted odds ratio of 1.21 (95% CI: 1.11 – 1.32, p < 0.0001). Similarly, key comorbidities including diabetes (aOR 1.32, p < 0.0001), hypertension (aOR 1.25, p < 0.0001), and high triglyceride levels (aOR 1.19, p = 0.0003) were significantly linked to increased depressive symptoms.

Education level and race/ethnicity were not strong predictors in the model. Although race/ethnicity reached statistical significance (p = 0.0175), the effect size was small, and education level did not show a significant association with depressive symptoms (p = 0.1472).

## Discussion

Our study highlights a statistically significant but modest association between VDD and depressive symptoms. After adjusting for demographic and clinical variables, VDD was associated with an 18% increase in odds of experiencing any depressive symptoms, supporting the hypothesis that vitamin D plays a contributory role in mental health. This finding is consistent with prior observational studies and meta-analyses (Anglin et al., 2013; dos Santos et al., 2024; Mulugeta et al., 2020), and reinforces the emerging view that low vitamin D status may influence mood regulation through several biological mechanisms.

In particular, individuals with VDD had a higher prevalence of specific symptoms such as low interest, feeling down, and low energy (Table 3), although suicidal ideation did not differ significantly between deficient and sufficient groups. These findings suggest that vitamin D may more strongly influence specific domains of depression, particularly little interest, low energy and depression, which have biological links to inflammation and neurotransmitter imbalance— both potentially modulated by vitamin D (Eyles et al., 2005; McGrath & Féron, 2004).

The demographic analysis (Table 1) revealed that VDD was more prevalent among younger adults and those with higher education, an unexpected finding that may reflect lifestyle patterns such as decreased sun exposure among educated individuals with predominantly indoor occupations (Mulugeta et al., 2020). Obesity was also notably more common in the VDD group, aligning with previous literature suggesting that adiposity may sequester vitamin D, reducing its bioavailability (Holick, 2007). Comorbidities such as diabetes and hypertension, though not significantly different in prevalence between groups, were strong predictors of depression in the multivariate model (Table 4), suggesting independent contributions to mental health burden (Raza et al., 2025).

**Table 4:**
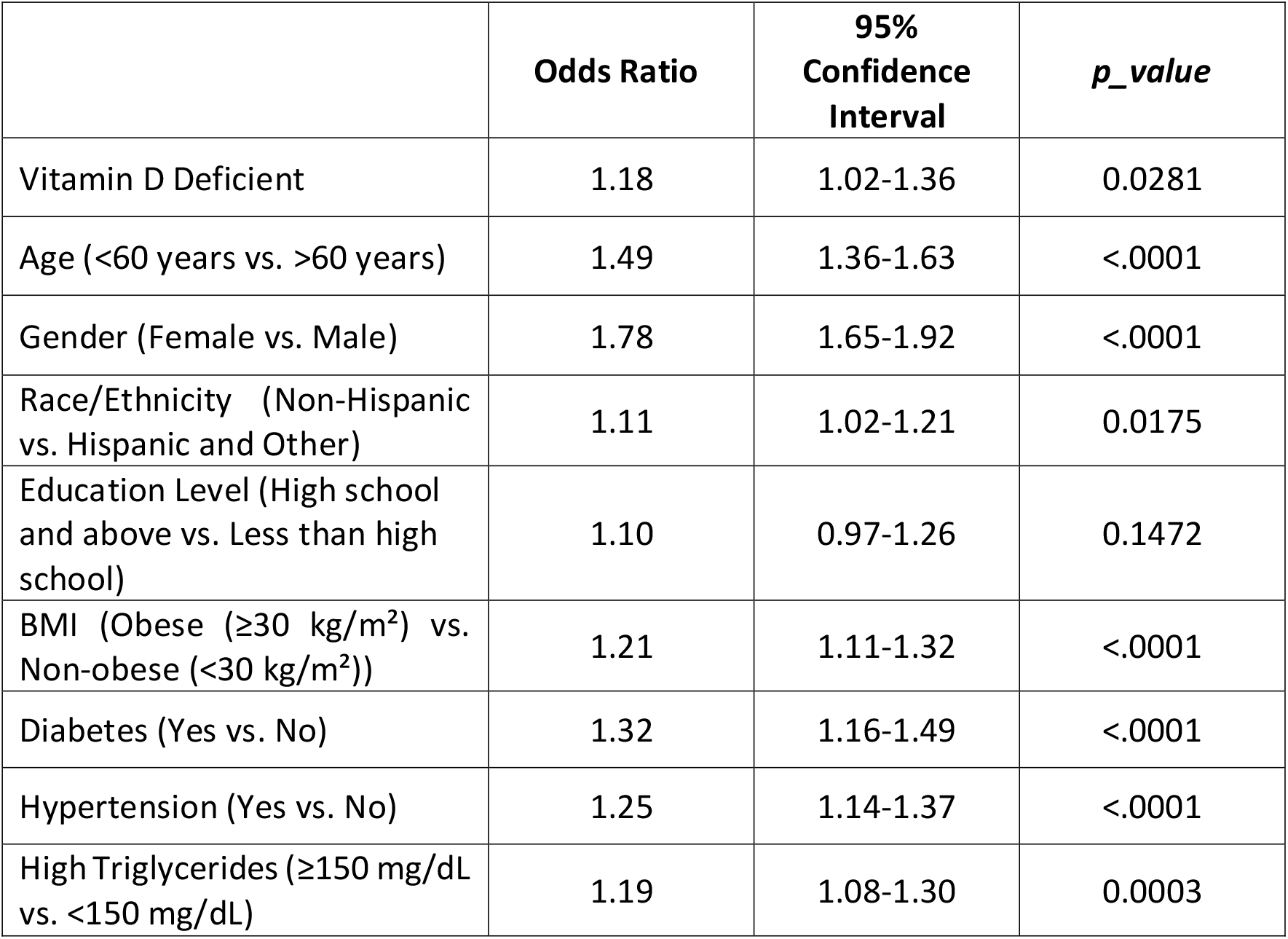
Multivariate analysis on predictors for having any depressive symptoms.

|  | **Odds Ratio** | **95%**<br>**Confidence**<br>**Interval** | ***p_value*** |
| --- | --- | --- | --- |
| Vitamin D Deficient | 1.18 | 1.02-1.36 | 0.0281 |
| Age (<60 years vs. >60 years) | 1.49 | 1.36-1.63 | <.0001 |
| Gender (Female vs. Male) | 1.78 | 1.65-1.92 | <.0001 |
| Race/Ethnicity (Non-Hispanic vs. Hispanic and Other) | 1.11 | 1.02-1.21 | 0.0175 |
| Education Level (High school and above vs. Less than high school) | 1.10 | 0.97-1.26 | 0.1472 |
| BMI (Obese ($\geq 30$ kg/m<sup>2</sup>) vs. Non-obese (<30 kg/m<sup>2</sup>)) | 1.21 | 1.11-1.32 | <.0001 |
| Diabetes (Yes vs. No) | 1.32 | 1.16-1.49 | <.0001 |
| Hypertension (Yes vs. No) | 1.25 | 1.14-1.37 | <.0001 |
| High Triglycerides ($\geq 150$ mg/dL vs. <150 mg/dL) | 1.19 | 1.08-1.30 | 0.0003 |

Gender differences in depression were striking, with females exhibiting nearly 78% higher odds of depressive symptoms—consistent with longstanding epidemiologic trends and biologic explanations including hormonal regulation and psychosocial stress exposure (Goodwin & Gotlib, 2004; Drevets et al., 2008; Malhi & Mann, 2018). Age was also a significant predictor, with individuals ≥60 years showing lower odds of depressive symptoms, a finding attributed in prior research to enhanced emotional regulation or cohort resilience (Fiske et al., 2009). Mechanistically, vitamin D may influence mood via its role in regulating serotonin synthesis, neuroplasticity, and immune modulation. The widespread distribution of vitamin D receptors (VDR) in mood-related brain regions, such as the prefrontal cortex and hippocampus, supports this hypothesis (Eyles et al., 2005). Moreover, animal studies suggest that VDD may promote neuroinflammation and impair neurogenesis, further implicating vitamin D in mood regulation (Féron et al., 2005; McGrath & Féron, 2004). Genetic variations in VDR have also been associated with susceptibility to psychiatric disorders, stressing the complex, multifactorial nature of this relationship (Handoko et al., 2006).

While our findings support an association between VDD and depression, the effect size remains modest. This suggests that vitamin D status likely interacts with other biological and environmental factors in a bi-directional manner—where depression may lead to behaviors that reduce sun exposure and dietary quality, and VDD may exacerbate mood disturbances (Kim et al., 2020; Mulugeta et al., 2020). This nuanced relationship underscores the importance of considering VDD as a modifiable risk factor rather than a sole causative agent.

This study is limited by its cross-sectional design, precluding causal inference. Self-reported depressive symptoms may be subject to bias. Residual confounding may persist despite adjustment.

## Conclusion

Vitamin D deficiency is modestly associated with increased odds of depressive symptoms, particularly anhedonia and fatigue. While the effect is statistically significant, it is less pronounced than other predictors such as gender, hypertension, and diabetes. These findings highlight the need for a multifaceted approach to depression prevention that includes addressing nutritional deficiencies alongside metabolic, psychosocial, and behavioral risk factors. Future research should investigate whether correcting vitamin D deficiency through supplementation can meaningfully improve depressive symptoms, particularly in high-risk subgroups. Additionally, screening and correcting VDD may serve as one component of holistic mental health strategies, particularly in high-risk populations.

## Data Availability

All data produced are available online at https://www.cdc.gov/nchs/nhanes/

https://www.cdc.gov/nchs/nhanes/

